# Do amyloid cerebral deposits influence the long-term post-stroke cognitive outcome? The IDEA3 study

**DOI:** 10.1101/2024.07.18.24310673

**Authors:** Olivier Godefroy, Niels Trinchard, Etienne Marchal, Chantal Lamy, Sandrine Canaple, Marc-Etienne Meyer, Martine Roussel, Frank A Wollenweber, the IDEA3 study group

## Abstract

**Background:** Although the presence of amyloid deposits is associated with a more severe cognitive status in stroke patients at baseline, its influence on the subsequent cognitive outcome has not been extensively assessed. The primary objective of the present study of the IDEA3 cohort was to determine the influence of amyloid PET status on the 5-year cognitive outcome.

**Methods:** The 91 stroke patients (ischemic stroke: 89%; hemorrhagic stroke: 11%) with florbetapir PET data at baseline (positive: n=14) underwent comprehensive clinical and cognitive assessments for 5 years after the PET scan.

**Results:** A survival analysis (mean post-stroke follow-up: 80.4 ±27 months) showed that the incidence of dementia was higher in the PET-positive patients (OR=5.89, 95% CI: 1.24-22.7, p=0.02), as was the incidence of cognitive impairment (OR=10, 95% CI: 1.9-52.3, p=0.003). A Cox regression analysis showed that the association between amyloid status and the incidences of dementia (p=0.006) and CI (p=0.04) was still significant after adjustment for age. Considering the overall prevalence at last follow-up in the whole study population (n=91 patients), PET positivity was associated with an elevated risk of post-stroke cognitive impairment (OR=6.25, 95%CI: 1.77-22, p=0.002) or dementia (OR= 6, 95%CI: 1.76-20.5, p=0.002). The final Rankin score did not differ according to PET status (p=0.3).

**Conclusions:** Our results demonstrated the major impact of amyloid deposition on the stroke outcome and emphasized the need for comprehensive etiologic work-up in patients with post-stroke cognitive impairment.

**Clinical Trial Registration:** NCT 02813434

## Introduction

Some degree of post-stroke cognitive impairment (PSCI) is observed in at least half of stroke survivors, with a mild neurocognitive disorder in two thirds of these cases and dementia in the remaining third^3^. Although PSCI is generally attributed to vascular lesions alone, amyloid PET studies have revealed that the prevalence of amyloid positivity is approximately 15-20% depending on the age and frequency of cognitive impairment in the study population^1,2,4–7^. The two largest studies showed that the amyloid burden is associated with a more severe cognitive status at baseline^1,2^. Accordingly, the fact that 30% to 38%^1,7^ of cases of PS dementia are PET-positive supports the conclusions of the pioneering studies^8–10.^

The influence of amyloidopathy on the post-stroke (PS) cognitive outcome has been assessed in one study only; 6 (19.4%) of the 31 patients with delayed dementia (between 7 and 36 months) were PET-positive^2,11^ and had a steeper decline in the screening test scores^11^.

The primary objective of the present study of the IDEA3 cohort was to determine the influence of amyloid PET status on incident dementia. The secondary objectives were to determine incident cognitive impairment, impairments in five cognitive domains (action speed, executive function, memory, language, and visuoconstructive abilities), and the effect of PET status on the final Rankin score.

## Methods

The study’s methods and results were reported in accordance with the “Strengthening the Reporting of Observational Studies in Epidemiology” guidelines. The data that support the study’s findings are available from the corresponding author upon reasonable request. The study was performed in accordance with institutional guidelines and was approved by the regional investigational review board and informed consent was obtained from patients (Comité de Protection de Personnes Nord-Ouest II, Amiens, France; reference: 2013/27, 11 July 2013).

### Population

The IDEA3 cohort’s inclusion criteria and baseline characteristics have been described in detail elsewhere^1^. Briefly, the main inclusion criteria were age 40 or over, command of the French language, hospitalization in our university medical center for an acute ischemic or hemorrhagic stroke, available imaging data, the presence of a reliable informant, at least one impaired cognitive score at the follow-up assessment, consent to participation, and no fulfilment of any of exclusion criteria^12,13^. The main exclusion criteria (see Supplemental Methods 1.1.) included conditions (other than stroke) known to affect cognition, contra-indication to MRI, and the absence of informed consent. With regard to their demographic and clinical characteristics, the 91 study participants were typical of a hospital-based population of stroke patients (Table 1).

**Table 1.** Characteristics of the study cohort, overall and by amyloid PET status. Data are expressed as the frequency (percentages), mean ± standard deviations, or medians [interquartile range], as appropriate.

|  |  | Total | Amyloid PET |  | *p* |
| --- | --- | --- | --- | --- | --- |
|  |  |  | positive | Negative |  |
| Acute phase | N | 91 | 14 | 77 |  |
| | Age | 63.3 ± 10.7 | 72.6 ± 6.0 | 61.6 ± 10.5 | 0.0001 |
|  | Male sex | 68.1 | 71.4 | 67.5 | 1 |
|  | Right handedness | 89.0 | 92.9 | 88.3 | 1 |
| | Education (years) | 10.5 ± 2.8 | 9.6 ± 1.7 | 10.6 ± 2.9 | 0.2 |
|  | Arterial hypertension | 61.5 | 64.3 | 61.0 | 1 |
|  | Hypercholesterolemia | 49.5 | 57.1 | 48.1 | 0.6 |
|  | Diabetes | 19.8 | 21.4 | 19.5 | 1 |
|  | Overweight | 70.3 | 64.3 | 71.9 | 0.4 |
|  | Current/ex- smoker | 20.9 / 16.5 | 28.6 / 14.3 | 19.4 / 16.9 | 0.4 / 1 |
|  | Current/previous alcohol abuse | 0.0 / 2.2 | 0.0 / 0.0 | 0.0 / 2.5 | 1 / 1 |
|  | History of atrial fibrillation | 18.7 | 35.7 | 15.6 | 0.13 |
|  | History of stroke/ischemic stroke | 5.5 / 3.3 | 14.3 / 14.3 | 3.9 / 1.3 | 0.17 / 0.6 |
| | Pre-stroke IQCODE score | 48.9 ± 2.5 | 52.3 ± 7.8 | 51.2 ± 8.0 | 0.6 |
|  | Pre-stroke Rankin score | 0 [0-0] | 0 [0-0] | 0 [0-1] | 0.13 |
| | Pre-stroke 4IADL score | 0.3 ± 1.1 | 0.86 ± 2.4 | 0.16 ± 0.5 | 0.4 |
| | NIHSS score on admission | 5.6 ± 5.8 | 4.7 ± 3.1 | 5.8 ± 6.2 | 0.3 |
|  | Acute complication | 37.7 | 30.0 | 39.0 | 0.7 |
|  | Delirium | 2.9 | 0.0 | 3.4 | 1 |
| Cause of stroke | Infarct subgroup (n = 81) | 89.0 | 85.7 | 89.6 | 0.6 |
|  | Atherosclerosis | 4.9 | 0.0 | 5.8 | 1 |
|  | Small vessel disease | 17.3 | 33.3 | 14.5 | 0.2 |
|  | Cardioembolic stroke | 24.7 | 33.3 | 23.2 | 0.5 |
|  | Other | 7.4 | 8.3 | 11.2 | 0.8 |
|  | Thrombolysis/Thrombectomy | 19.8 / 6.3 | 33.3 / 16.6 | 17.4 / 4.4 | 0.2 / 0.16 |
|  | Hemorrhage subgroup (n = 10) | 11.0 | 14.3 | 10.4 |  |
|  | Hypertensive | 50.0 | 0.0 | 75 | 0.13 |
|  | Amyloid angiopathy | 30.0 | 100 | 0 | 0.02 |
|  | Other | 20.0 | 0.0 | 25.0 | 1 |
| Baseline | Time post-stroke (days) | 808 ± 589 | 660 ± 515 | 835 ± 601 | 0.3 |
|  | Recurrent stroke | 1.1 | 0.0 | 1.3 | 1 |
| | NIHSS score | 1.49 ± 2.02 | 0.86 ± 2.4 | 0.16 ± 0.5 | 0.8 |
| | Depressive symptoms | 14.5 ± 11.3 | 15.7 ± 11.1 | 14.3 ± 11.3 | 0.7 |
| | MMSEa score | 25.9 ± 3.6 | 23.1 ± 4.8 | 26.4 ± 3.1 | 0.023 |
| | MoCA score | 22.3 ± 4.4 | 16.7 ± 4.9 | 22.7 ± 4.2 | 0.027 |
| | 4IADL score | 1.76 ± 2.31 | 2.0 ± 3.16 | 1.71 ± 2.14 | 0.7 |
|  | Rankin score | 2 [1-3] | 2 [1-3] | 2 [1-3] | 0.9 |

IQCODE: Informant Questionnaire on Cognitive Decline in the Elderly, 4IADL: score for 4 instrumental activities of daily living, NIHSS: National Institute of Health Stroke Scale, MMSEa: education-adjusted score on the MiniMental Status Examination, MoCA: Montréal Cognitive assessment.

### Pre-inclusion and baseline assessments

As detailed elsewhere^1^, the pre-inclusion visit was scheduled at six months PS and included clinical and neuropsychological assessments. The baseline visit (M0) corresponded to the day of the PET examination and included clinical and neuropsychological assessments and MRI.

### PET assessment

The amyloid PET scan (performed in accordance with current guidelines, see Supplement Methods 3) was positive in 14 of the 91 patients in the IDEA3 cohort. Due to delays in the delivery of florbetapir, the mean time interval between the stroke and the amyloid PET scan was approximately two years and did not differ according to PET status (660 ± 515 days in the PET-positive group and 835 ± 601 days in the PET-negative group; p=0.3)^1^.

### Clinical and cognitive assessments

The clinical and cognitive assessments have been detailed elsewhere^12,13^ (see Supplement Methods 2). Briefly, the clinical assessment gathered data on new medical events, current treatments, neurological impairment using the National Institute of Health Stroke Scale (NIHSS) score^14^, gait score^15^, the functional outcome (on the Rankin scale, graded with a structured interview^16^), the Barthel index^17^ and the Instrumental Activities of Daily Living (IADL) scale^15,18^. The neuropsychological assessments included the Mini Mental Status Examination^19,20^, the Montreal Cognitive Assessment^21^, and the GRECogVASC comprehensive battery^12,13,22^.

At baseline, cognitive impairment was observed in 25 patients: 17 had mild cognitive impairment (15 [19.5%] in the PET-negative subgroup and 2 [14.3%] in the PET-positive subgroup), and 8 had dementia (6 [7.8%] in the PET-negative subgroup and 2 [14.3%] in the PET-positive subgroup).

### Follow-up assessments

Annual neurological and cognitive follow-up visits were scheduled up until five years after the inclusion visit (i.e. from M12 to M60). The present longitudinal study focused on data collected after the baseline visit. If the patient could not attend the visit (especially during the coronavirus disease 2019 pandemic), a telephone interview was organized. During the telephone follow-up interview, only the following variables were documented: the occurrence of a new medical event, current treatments, the gait score^23^, cognitive status (estimated using the AD8 and with 2 as the cutoff score)^24^, the functional outcome (using the Rankin scale^16^, the Barthel index^17^ and the Instrumental Activities of Daily Living scale^15,18^).

### Statistical analysis

The association between the 5-year cognitive outcomes and amyloid status was analyzed in a log-rank test^25^. The primary outcome was the presence of dementia. The secondary outcomes were presence of cognitive impairment (regardless of its severity, i.e. mild or severe) and impairments in the five cognitive domains (action speed, executive functions, episodic memory, language and visuoconstructive abilities). The time interval was expressed as the time PS.

A Cox regression analysis with stepwise factor selection^26^ was used to examine the influence of the following confounding factors: age, educational level, prestroke Rankin score^16^, prestroke restriction of IADL^13^, prestroke cognitive status (examined using the Informant Questionnaire on Cognitive Decline in the Elderly (IQCode) score^27^), stroke type (ischemic or hemorrhagic), PET status, and the PET status x stroke type interaction. Factors that were significant in the bivariate step were submitted to a Cox regression analysis. The influence of PET status on the final Rankin score was examined using a Mann-Whitney test. All statistical analyses were performed using SAS software (SAS Institute, Inc., Cary, NC). The threshold for statistical significance was set to p≤0.05, unless otherwise indicated.

## Results

Of the 91 included patients, 74 (81.3%) completed the M60 visit, and 53 (58.2%) completed the M60 cognitive battery (Supplemental Table 1). Twenty-two (24.2%) patients were assessed over the telephone: 19 (24.7%) in the PET-negative group and 3 (21.4%) in the PET-positive group (p=0.8). The mean follow-up duration did not differ (p=0.16) according to PET status (positive: 70.7 ±32.7 months; negative: 82.2 ±26.8 months). Five patients (4 in the PET-negative group) died during the study period; of these, two had dementia and three had subjective cognitive decline at the last follow-up visit.

The survival analysis showed that the incidence of dementia was higher in the PET-positive group (p=0.02) (Figure 1): in the 83 patients without dementia at baseline, incident dementia was observed in 4 (51.1%) of the 7 PET-positive patients and 7 (11.8%) of the 65 PET-negative patients (odds ratio [OR]=5.89, 95% confidence interval [CI]: 1.24-22.7, p=0.02).

**Figure 1.**
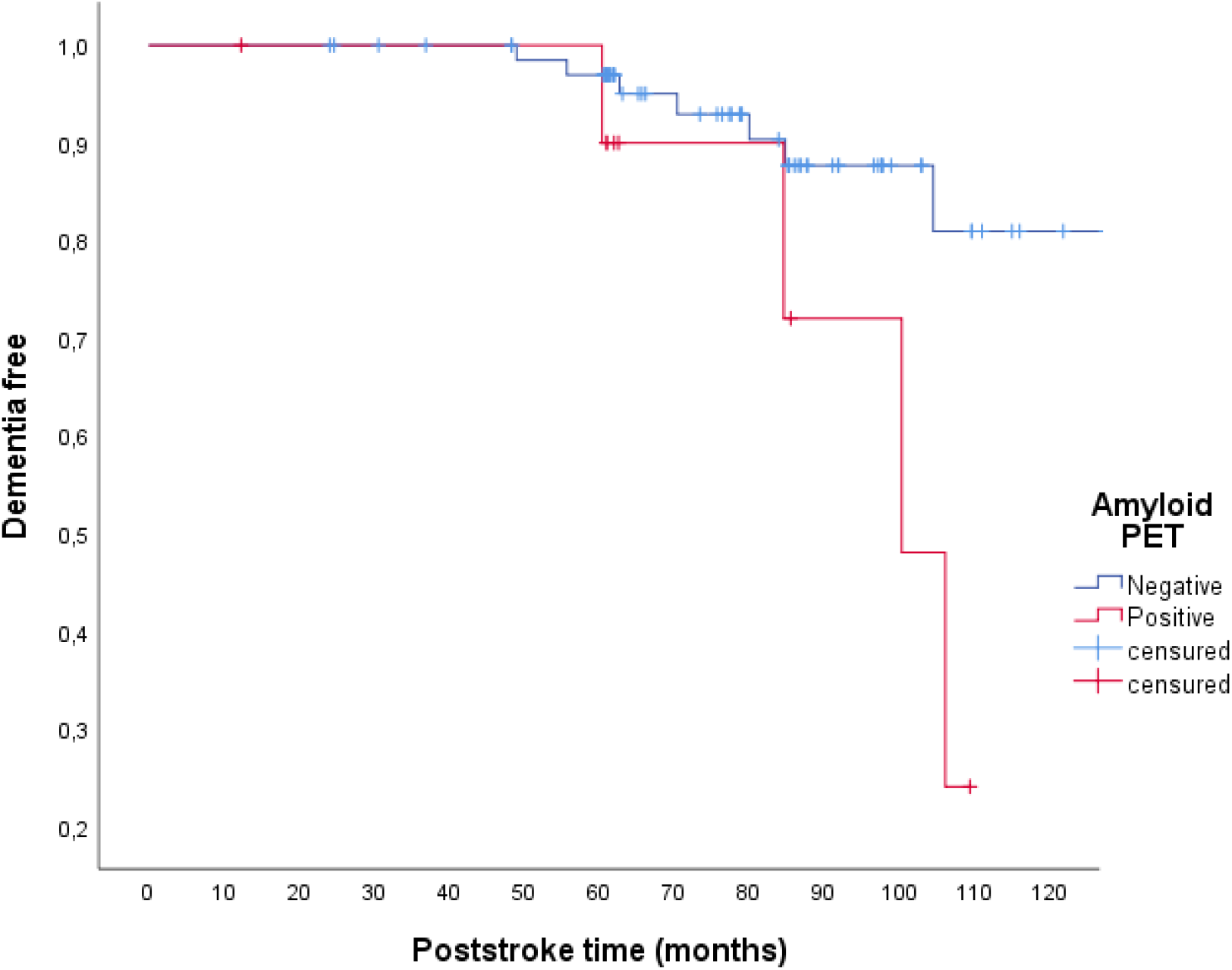
The risk of developing incident dementia.

Among the variables fed into the Cox regression analysis, the bivariate step showed that PET positivity (p=0.006), age (p=0.014), and PET status x stroke type interaction (p=0.03) were associated with incident cognitive impairment (educational level: p=0.9; prestroke Rankin score: p=0.4; prestroke IADL: p=0.4; IQCode: p=0.8; stroke type: p=0.8). In the multivariate step, only PET status was significant (p=0.006).

The survival analysis showed that the incidence of cognitive impairment was higher in the PET-positive group (p=0.007) (Figure 2): in the 66 patients without cognitive impairment at baseline, incident cognitive impairment was observed in 8 (80%) of the 10 PET-positive patients and 16 (28.6%) of the 56 PET-negative patients (OR=10, 95% CI: 1.9-52.3, p=0.003).

**Figure 2.**
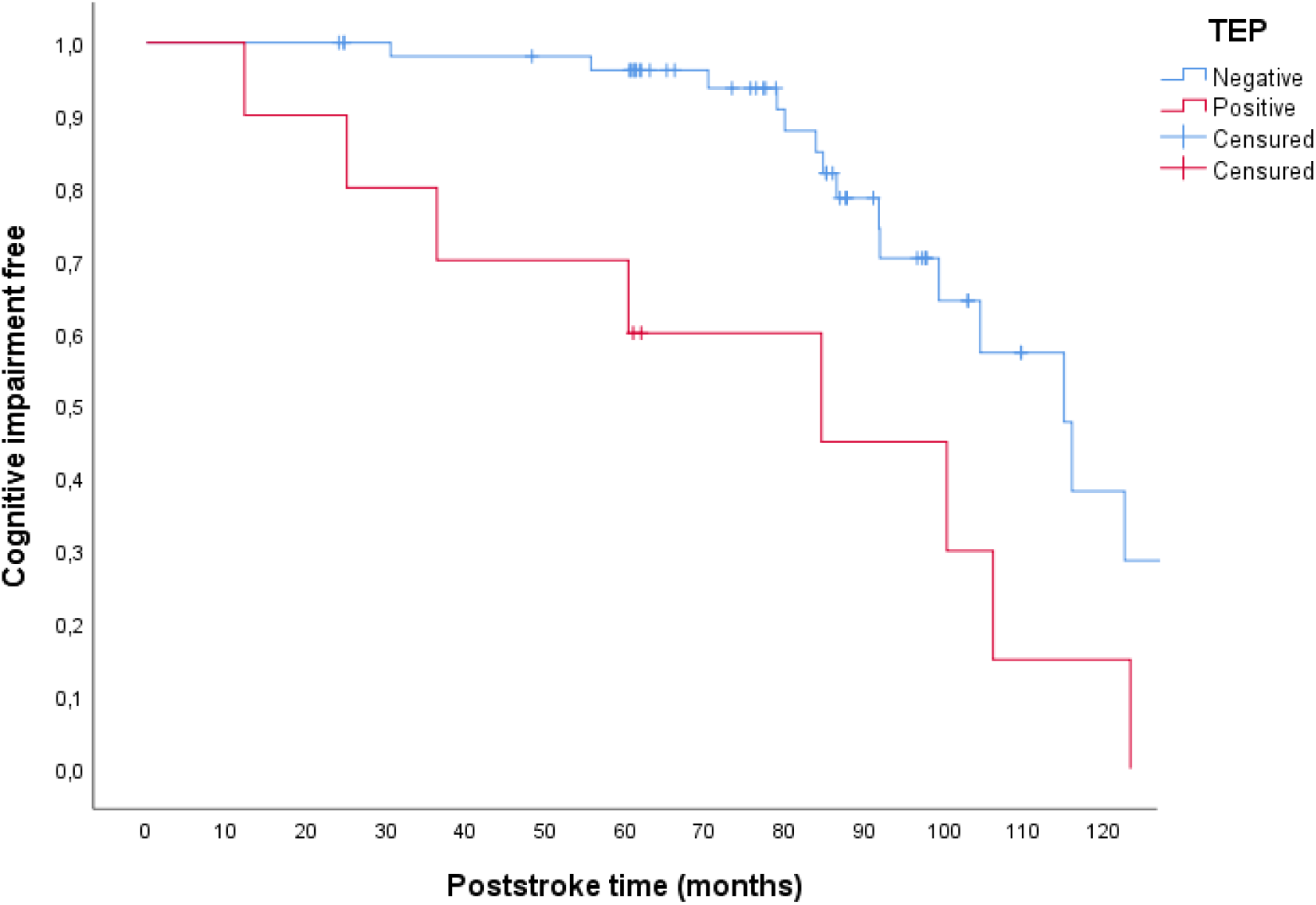
The risk of developing incident cognitive impairment.

Among the variables submitted to Cox regression analysis, the bivariate step showed that PET positivity (p=0.02) and age (p=0.035) were significantly associated with incident dementia (education: p=0.13; prestroke Rankin score: p=0.08; prestroke IADL: p=0.2; IQCode: p=0.8; stroke type: p=0.6; PET status x stroke type: p=0.2). In the multivariate step, only PET status was significant (p=0.044).

Considering overall prevalence at last follow-up in the whole population of 91 patients, PET positivity was associated with an elevated risk of PSCI (PET-positive: n=10 [71.4%]; PET-negative: n=22 [28.6%]; OR=6.25, 95%CI: 1.77-22, p=0.002) and an elevated risk of dementia (PET-positive: n=7 [50%]; PET-negative: n=11 [14.3%]; OR= 6, 95%CI: 1.76-20.5, p=0.002).

Considering the secondary outcomes analyzed in the 66 patients without overall cognitive impairment at baseline, PET positivity was associated with a higher probability of developing an impairment at last follow-up in episodic memory (p=0.002) and language (p=0.02) (Supplementary Figures; action speed: p=0.4; executive functions: p=0.2; visuoconstructive abilities: p=0.3).

The final Rankin score did not vary according to the PET status (p=0.3), although there was a higher proportion of scores of >3 in the PET-positive group (n=5 [35.7%]) than in the PET-negative group (12 [2.6%]).

## Discussion

Our longitudinal study of the IDEA3 cohort showed that cerebral amyloid PET positivity is associated with (i) a fivefold greater risk of developing incident dementia, (ii) a sixfold greater risk of developing PSCI and PS dementia; and (iii) more pronounced cognitive deterioration in language and memory. The greater risk of developing dementia was independent of age, educational level, prestroke status, and stroke type. These results extend the previous report of a higher frequency of PET positivity in stroke patients experiencing delayed dementia (within 3 years of stroke)^2^; in our comprehensive cognitive assessment of a larger population after a mean follow-up period of 6.7 years, the risk of a poor cognitive outcome increased by a factor of 5 to 6. The fact that the interaction with the stroke subtype was not significant indicates that the risk of a poor cognitive outcome was associated with amyloid deposition in general (i.e. regardless its clinical presentation and the presence of Alzheimer disease or amyloid angiopathy^28–31^).

Our study had several limitations. Firstly, the delayed delivery of florbetapir meant that the PET examination was not performed in the first year PS for 59 patients. However, the amyloid burden was not associated with the time interval between the stroke and the PET scan. This finding is consistent with the very long time course of amyloid deposition (i.e. detection of amyloid 15 years before symptom onset in Alzheimer disease)^32^ and the stroke’s lack of influence on amyloid deposition^1,4,33^. Thus, the two-year time interval between stroke and the PET scan does not preclude an analysis of the impact of amyloid status on subsequent cognitive impairment. Secondly, the small sample size in the PET-positive group (n = 14) limited the study’s power. Again, this sample size did not preclude relevant findings, and our cohort is the largest cohort yet studied.

In conclusion, the results of the present study demonstrated the major impact of amyloid deposition on stroke outcome. Our findings emphasize the need for comprehensive etiologic work-up in patients with PSCI. Further research is needed to identify the characteristics of at-risk stroke patients who should be specifically screened for Alzheimer disease.

## Data Availability

The data that support the study?s findings are available from the corresponding author upon reasonable request.

## List of Non-standard Abbreviations

PS: poststroke
CI: cognitive impairment
NIHSS: National Institute of Health Stroke Scale
IADL: Instrumental Activities of Daily Living
IDEA3: Imagerie des dépôts amyloïdes cérébraux par florbetapir AV-45 et diagnostic des déficits cognitifs et démence post Accident Vasculaire Cérébral

## Acknowledgments

We thank Hassan Berrissoul and Astrid Causel for assistance with the organizational aspects of the study and collecting clinical data, as well as Quentin Legendre for help in collecting the MRI data.

## References

1. Godefroy O, Barbay M, Martin J, et al. Prevalence of Amyloid Cerebral Deposits and Cognitive Outcome After Stroke: The IDEA3 Study. J Stroke. 2023;25(2):315–319. doi:10.5853/jos.2022.03391

2. Mok VCT, Lam BYK, Wang Z, et al. Delayed-onset dementia after stroke or transient ischemic attack. Alzheimers Dement J Alzheimers Assoc. 2016;12(11):1167–1176. doi:10.1016/j.jalz.2016.05.007

3. Barbay M, Diouf M, Roussel M, Godefroy O, GRECOGVASC study group. Systematic Review and Meta-Analysis of Prevalence in Post-Stroke Neurocognitive Disorders in Hospital-Based Studies. Dement Geriatr Cogn Disord. 2018;46(5-6):322–334. doi:10.1159/000492920

4. Wollenweber FA, Därr S, Müller C, et al. Prevalence of Amyloid Positron Emission Tomographic Positivity in Poststroke Mild Cognitive Impairment. Stroke. 2016;47(10):2645–2648. doi:10.1161/STROKEAHA.116.013778

5. Hagberg G, Ihle-Hansen H, Fure B, et al. No evidence for amyloid pathology as a key mediator of neurodegeneration post-stroke - a seven-year follow-up study. BMC Neurol. 2020;20(1):174. doi:10.1186/s12883-020-01753-w

6. Koenig LN, McCue LM, Grant E, et al. Lack of association between acute stroke, post-stroke dementia, race, and β-amyloid status. NeuroImage Clin. 2021;29:102553. doi:10.1016/j.nicl.2020.102553

7. Yang J, Wong A, Wang Z, et al. Risk factors for incident dementia after stroke and transient ischemic attack. Alzheimers Dement. 2015;11(1):16–23. doi:10.1016/j.jalz.2014.01.003

8. Tatemichi TK, Foulkes MA, Mohr JP, et al. Dementia in stroke survivors in the Stroke Data Bank cohort. Prevalence, incidence, risk factors, and computed tomographic findings. Stroke. 1990;21(6):858–866. doi:10.1161/01.str.21.6.858

9. Kokmen E, Whisnant JP, O’Fallon WM, Chu CP, Beard CM. Dementia after ischemic stroke: a population-based study in Rochester, Minnesota (1960-1984). Neurology. 1996;46(1):154–159. doi:10.1212/wnl.46.1.154

10. Hénon H, Durieu I, Guerouaou D, Lebert F, Pasquier F, Leys D. Poststroke dementia: incidence and relationship to prestroke cognitive decline. Neurology. 2001;57(7):1216–1222. doi:10.1212/wnl.57.7.1216

11. Liu W, Wong A, Au L, et al. Influence of Amyloid-β on Cognitive Decline After Stroke/Transient Ischemic Attack: Three-Year Longitudinal Study. Stroke. 2015;46(11):3074–3080. doi:10.1161/STROKEAHA.115.010449

12. Godefroy O, Leclercq C, Roussel M, et al. French adaptation of the vascular cognitive impairment harmonization standards: the GRECOG-VASC study. Int J Stroke Off J Int Stroke Soc. 2012;7(4):362–363. doi:10.1111/j.1747-4949.2012.00794.x

13. Barbay M, Taillia H, Nédélec-Ciceri C, et al. Prevalence of Poststroke Neurocognitive Disorders Using National Institute of Neurological Disorders and Stroke-Canadian Stroke Network, VASCOG Criteria (Vascular Behavioral and Cognitive Disorders), and Optimized Criteria of Cognitive Deficit. Stroke. 2018;49(5):1141–1147. doi:10.1161/STROKEAHA.117.018889

14. Brott T, Adams HP, Olinger CP, et al. Measurements of acute cerebral infarction: a clinical examination scale. Stroke. 1989;20(7):864–870. doi:10.1161/01.str.20.7.864

15. Tasseel-Ponche S, Barbay M, Roussel M, et al. Determinants of disability at 6 months after stroke: The GRECogVASC Study. Eur J Neurol. 2022;29(7):1972–1982. doi:10.1111/ene.15319

16. Godefroy O, Just A, Ghitu A, et al. The Rankin scale with revised structured interview: effect on reliability, grading of disability and detection of dementia. Int J Stroke Off J Int Stroke Soc. 2012;7(2):183. doi:10.1111/j.1747-4949.2011.00743.x

17. Mahoney FI, Barthel DW. FUNCTIONAL EVALUATION: THE BARTHEL INDEX. Md State Med J. 1965;14:61–65.

18. Lawton MP, Brody EM. Assessment of older people: self-maintaining and instrumental activities of daily living. The Gerontologist. 1969;9(3):179–186.

19. Folstein MF, Folstein SE, McHugh PR. “Mini-mental state”. A practical method for grading the cognitive state of patients for the clinician. J Psychiatr Res. 1975;12(3):189–198. doi:10.1016/0022-3956(75)90026-6

20. Kalafat M, Hugonot-Diener L, Poitrenaud J. Standardisation et étalonnage français du “Mini Mental State” (MMS) version GRÉCO. [French standardization and range for the GRECO version of the “Mini Mental State” (MMS).]. Rev Neuropsychol. 2003;13(2):209–236.

21. Nasreddine ZS, Phillips NA, Bédirian V, et al. The Montreal Cognitive Assessment, MoCA: a brief screening tool for mild cognitive impairment. J Am Geriatr Soc. 2005;53(4):695–699. doi:10.1111/j.1532-5415.2005.53221.x

22. Roussel M, Godefroy O. La batterie GRECOGVASC: Evaluation et diagnostic des troubles neurocognitifs vasculaires avec ou sans contexte d’accident vasculaire cérébral. De Boeck Superieur; 2016.

23. Garcia PY, Roussel M, Bugnicourt JM, et al. Cognitive impairment and dementia after intracerebral hemorrhage: a cross-sectional study of a hospital-based series. J Stroke Cerebrovasc Dis Off J Natl Stroke Assoc. 2013;22(1):80–86. doi:10.1016/j.jstrokecerebrovasdis.2011.06.013

24. Galvin JE, Roe CM, Powlishta KK, et al. The AD8: A brief informant interview to detect dementia. Neurology. 2005;65(4):559–564. doi:10.1212/01.wnl.0000172958.95282.2a

25. Mantel N. Evaluation of survival data and two new rank order statistics arising in its consideration. Cancer Chemother Rep. 1966;50(3):163–170.

26. Cox DR. Some Statistical Methods Connected with Series of Events. J R Stat Soc Ser B Methodol. 1955;17(2):129–157. doi:10.1111/j.2517-6161.1955.tb00188.x

27. Jorm AF. A short form of the Informant Questionnaire on Cognitive Decline in the Elderly (IQCODE): development and cross-validation. Psychol Med. 1994;24(1):145–153. doi:10.1017/s003329170002691x

28. Clark CM, Schneider JA, Bedell BJ, et al. Use of florbetapir-PET for imaging beta-amyloid pathology. JAMA. 2011;305(3):275–283. doi:10.1001/jama.2010.2008

29. Dugger BN, Clark CM, Serrano G, et al. Neuropathologic heterogeneity does not impair florbetapir-positron emission tomography postmortem correlates. J Neuropathol Exp Neurol. 2014;73(1):72–80. doi:10.1097/NEN.0000000000000028

30. Planton M, Saint-Aubert L, Raposo N, et al. Florbetapir Regional Distribution in Cerebral Amyloid Angiopathy and Alzheimer’s Disease: A PET Study. J Alzheimers Dis JAD. 2020;73(4):1607–1614. doi:10.3233/JAD-190625

31. Gurol ME, Becker JA, Fotiadis P, et al. Florbetapir-PET to diagnose cerebral amyloid angiopathy. Neurology. 2016;87(19):2043–2049. doi:10.1212/WNL.0000000000003197

32. Bateman RJ, Xiong C, Benzinger TLS, et al. Clinical and biomarker changes in dominantly inherited Alzheimer’s disease. N Engl J Med. 2012;367(9):795–804. doi:10.1056/NEJMoa1202753

33. Sahathevan R, Linden T, Villemagne VL, et al. Positron Emission Tomographic Imaging in Stroke: Cross-Sectional and Follow-Up Assessment of Amyloid in Ischemic Stroke. Stroke. 2016;47(1):113–119. doi:10.1161/STROKEAHA.115.010528

